# Associations of plasma ergothioneine levels with cognitive function change in non-demented older Chinese adults: A community-based longitudinal study

**DOI:** 10.1101/2025.07.16.25331363

**Authors:** Danyang Zhao, Chengwu Feng, Peiyu Li, Yunan Liu, Long Lu, Bin Jiang, Shifu Xiao, Geng Zong

**Author notes:** Shifu Xiao and Geng Zong contributed equally as senior authors. Correspondence to: Shifu Xiao, MD, Department of Geriatric Psychiatry, Shanghai Mental Health Center, Shanghai Jiao Tong University School of Medicine, 600 South Wanping Road, Shanghai 200032, China; Alzheimer’s Disease and Related Disorders Center, Shanghai Jiao Tong University, 600 South Wanping Road, Shanghai 200032, China. Email, Geng Zong, PhD, Institute of Nutrition, Fudan University, No.130 Dong-An Road, Shanghai 200032, China. Email.

## Abstract

**Aim:** Animal studies have reported that ergothioneine (ET) exert a protective effect on cognitive function, but evidence from humans remains limited. In this study, we explored the association between plasma ET levels and cognitive change in a prospective cohort study specifically focused on older adults without dementia in Shanghai.

**Methods:** This analysis was based on 1,131 community-dwelling elder Chinese (mean age 69 years) from the Shanghai Brain Aging Study. Ultra-high-performance liquid chromatography-mass spectrometry was used to measure plasma ET at baseline. Cognitive function was assessed at baseline and follow-up (mean 2.0 years) via the Montreal Cognitive Assessment (MoCA) by trained clinicians. Multivariable linear regression evaluated ET-cognition associations with confounder adjustment.

**Results:** Each 1-SD increase in ET was associated with a 0.23 higher point in MoCA changes (95% CI: 0.04–0.43, *P*=0.021) after adjustment for age, sex, smoking, alcohol, diet score, educational level, BMI, hypertension, hyperlipidemia, diabetes, cardiovascular disease, *APOE ε4* status and baseline MoCA scores. Sex-stratified analysis revealed a significant association in men (*β*=0.78 per SD, 95% CI: 0.35–1.21, *P* < 0.001), but not in women (*P* _for interaction_ = 0.017). Dose-response curves indicated plateauing effects above 1000 ng/mL. Domain-specific analyses identified significant protective associations for visuospatial/executive function (*β*=0.08, 95% CI: 0.02–0.14) and delayed recall (*β*=0.11, 95% CI: 0.02–0.20).

**Conclusions:** Our findings indicate that higher plasma ET levels are significantly associated with slower cognitive decline independent of confounders in non-demented community-dwelling elderly participants, with such association observed in men but not women.

## Background

An estimated 57 million people worldwide currently suffer from dementia in 2019, with projections suggesting a tripling of this number by 2050^1^. Given the central roles of oxidative stress and neuroinflammation in dementia pathogenesis, dietary antioxidants have emerged as promising candidates for cognitive preservation^2^.

Ergothioneine (ET), a diet-derived sulfur-containing amino acid derivative and potent thione-based antioxidant, is primarily obtained from mushrooms and fermented foods^3^. Its absorption and tissue distribution in humans are mediated by the organic cation transporter OCTN1 (encoded by *SLC22A4*), which facilitates pH-dependent intestinal uptake^4^ and selective accumulation in organs^5^. Notably, ET has been detected in human parietal brain tissues^6^ and medulla oblongata^7^, confirming its ability to cross the blood-brain barrier. Animal studies have shown that ET may protect against neuronal damage induced by cisplatin^8^, β-amyloid (Aβ)^9,10^, and age-related cognitive decline^11^. Emerging observational studies have found that lower plasma levels of ET are significantly associated with higher risks of neurodegenerative diseases, such as mild cognitive impairment (MCI)^12,13^ and dementia^14–18^, however, several knowledge gaps remain. First, most of the above studies were based on cross-sectional study design^12–17^, therefore potential reverse causation cannot be excluded. It has been suggested that plasma ET declines concomitantly with the deterioration of cognitive function^14^. Second, since the impact of a single dietary factor on health is mild, it is prone to be affected by the baseline characteristics of subjects (such as sex, educational level, disease status and gene polymorphism). However, no study has systematically evaluated potential effect modifiers on the association between ET levels and cognitive function. Third, the dose-response distribution between ET and cognitive function remains undetermined.

To fill these gaps, we utilized prospectively collected data of 1,131 community-dwelling older adults from the Shanghai Brain Aging Study (SBAS) cohort, and examined the association between baseline plasma ET levels and subsequent changes in cognitive function. We particularly analyzed the potential effect modification of baseline characteristics (e.g., sex, educational level, disease status and genetic factor) on the associations and explore the dose-response associations between ET and cognitive decline.

## Methods

### Study population

The Shanghai Brain Aging Study (SBAS) is an ongoing population-based prospective cohort study established in 2016 to investigate the influencing factors related to cognitive decline among community-dwelling adults aged ≥55 years in Shanghai, China. Follow-up assessments have been conducted biennially since baseline, with three waves completed as of 2024. At enrollment, a total of 2,430 eligible residents were screened, of whom 1,627 participants: (1) completed standardized in-person interviews, (2) provided fasting blood specimens at baseline, and (3) authorized health insurance data linkage, thus included for follow-up. The study protocol was approved by the Shanghai Clinical Research Ethics Committee (Approval No. 2018-10-16), and all participants provided written informed consent.

For the current analysis, baseline and first follow-up data (mean follow-up duration: 2.0 ± 0.1 years; retention rate: 74.0%) were used to minimize sample reduction because of death and loss to follow-up. We excluded 88 participants with dementia at baseline and an additional 408 participants with missing data on the Montreal Cognitive Assessment (MoCA) at either baseline or first follow-up, resulting in a final sample of 1,131 participants (eFigure 1).

### Cognitive assessments

Cognitive function was assessed using the MoCA (Beijing Version) following standardized protocols at baseline and during each follow-up visit^19,20^. All assessments were administered by trained clinicians. The MoCA domains include visuospatial/executive, naming, memory, attention, language, abstraction, delayed recall and orientation. The range of overall score was between 0 and 30 points. The primary outcome was defined as the absolute change in MoCA scores during follow-up (ΔMoCA = MoCA _follow-up1_ –MoCA _baseline_).

### Plasma Ergothioneine Measurements

At each research site, fasting blood samples (10 mL) were collected in EDTA tubes from participants after a 12-hour overnight fast in the following morning. All samples were centrifuged at 4°C and stored at −80°C until analysis. Informed consent was signed by all participants prior to sample collection. An internal standard (IS) solution was prepared in methanol:acetonitrile (1:1, v/v) containing: L-glutamic acid-2,3,3,4,4-d5 (2.5 μg/mL). A stock solution (5 mg/mL) was prepared by dissolving ET standard in 50% methanol.

A randomized sequence was employed for continuous sample analysis, with a quality control sample added to each batch of samples (n = 10) during instrumental analysis. The analysis was performed as described by Luo et al^21^. Samples were separated using the Vanquish ultra-high-performance liquid chromatography system (Agilent 1290 Infinity III) and then subjected to mass spectrometry with a Q Exactive HF-X mass spectrometer (Thermo). Details of sample pretreatment and instrumental parameters were provided in the **Supplementary Method.**

Chromatographic peak integration for ET and calibration curve construction (prepared by gradient dilution of the stock solution) were performed using TraceFinder software, with quantification achieved via IS method. The linear calibration curve for ET was y = 0.0001041x - 0.002337 with an R² of 0.9951 and a linear range of 50–25000 ng/mL, while the coefficients of variation for quality control samples were 19.6%.

### Ascertainment of Covariates

A structured questionnaire was administered by trained interviewers to collect demographic characteristics including age, sex, and highest attained education. Educational level was categorized as illiterate, primary school, middle school and high school or above. Smoking status was dichotomized as “ever-smoker” (current or former smoking) versus “never-smoker”, while alcohol use was defined as “ever-drinker” versus “never-drinker”. Dietary quality was evaluated using a composite diet score (range 0-4) based on four components: 1 point each for daily intake of fruits, vegetables, or fish, and 1 point for abstaining from red meat consumption. Scores were summed to create an ordinal diet quality metric.

Anthropometric measurements including body weight, height and blood pressure were obtained following standardized protocols. Body mass index (BMI) was calculated as weight (kg)/height squared (m^2^). The history of diseases was ascertained from clinical interviews and health insurance data, with conditions classified as present or absent. Hypertension was defined as systolic blood pressure ≥140 mm Hg and/or diastolic blood pressure ≥90 mm Hg, or history of antihypertensive medication use. Diabetes mellitus was defined as fasting plasma glucose of 7.0 mmol/L or higher, or hypoglycemic medication use. Hyperlipidemia was defined as meeting any of the following criteria^22^: fasting plasma total cholesterol ≥6.2 mmol/L, LDL-C ≥4.1 mmol/L, triglycerides ≥2.3 mmol/L, HDL-C <1.0 mmol/L, or lipid-lowering medication use. Cardiovascular disease (CVD), specifically including stroke and coronary heart disease, was defined as clinical interviews or health insurance data.

CVD cases identified through health insurance data were defined using ICD-10 codes I20-I25 and I60-I64. Apolipoprotein E (*APOE*) genotyping was performed as described previously. Participants were categorized into three groups: *APOE ε4* carriers (defined as *ε3/ε4* or *ε4/ε4*), *APOE ε4* non-carriers and *APOE ε2/ε4* carriers.^23^.

### Statistical analysis

Plasma ET was normalized using the SD as the scaling factor^24^, when it was analyzed as a continuous variable (ng/mL per 1 SD). Owing to gender differences in plasma ET levels, sex-specific quartiles were derived for categorical analysis. When baseline characteristics were presented, mean and SD were reported for continuous variables, while n (percentages) was provided for categorical variables. We used one-way ANOVA to assess baseline differences in continuous variables across sex-stratified quartiles of ET, and χ^2^ tests for categorical values.

The associations of plasma ET concentrations with MoCA score change were explored using generalized linear models (GLM) with gaussian distribution and identity link function. We imputed missing values using means or modes if missingness is <5% of covariates, otherwise missing indicators were used. Plasma ET concentration was analyzed as a continuous variable (z-scaled) and sex-stratified quartiles. Three models of increasing complexity were designed. Model 1 was adjusted for age, sex and follow-up years. Model 2 was further adjusted for smoking, alcohol, diet score, educational level, BMI, *APOE ε4* status, baseline hypertension, hyperlipidemia, diabetes and cardiovascular disease. Model 3 was additionally adjusted for baseline MoCA scores. The same categories and adjustment models were used to assess MoCA domain score as the outcome. The linear trend was assessed by replacing categorical plasma ET levels with their median values in GLMs.

To assess the impact of covariates on the association between ET levels and cognitive function, we conducted the following two analyses. We conducted the partial correlation analysis between ET concentrations and all candidate covariates. Second, we performed a multivariable linear regression where the ET levels were treated as the outcome variable and regressed against all baseline phenotypes present.

We also used restricted cubic splines with three knots to explore the shape of the dose–response relationship between baseline plasma ET and changes in MoCA score. Additionally, the joint significance of the non-linear spline terms was evaluated through the ‘anova’ function implemented in the ‘rms’ R package, which tests the null hypothesis that all non-linear spline terms collectively equal zero. This approach avoids manual model comparisons by directly evaluating the contribution of non-linearity within the spline framework. These spline models were adjusted for the same potential confounders as the main multivariable linear regression analyses.

We conducted stratified analyses by sex (male vs. female), baseline MoCA score (median cutoff: <22 vs. ≥22), educational level (less than high school vs. high school or above), *APOE ε4* status (*APOE-ε4* carriers vs. *APOE-ε4* non-carriers) and hyperlipidemia (yes vs. no) as a priori defined and assessed potential interactions with these variables using the likelihood ratio test comparing two nested models: a full model containing the interaction term between the stratifying variable (e.g., sex) and plasma ET, and a reduced model excluding the interaction term. A *P*-value < 0.05 for the likelihood ratio chi-squared statistic indicated significant effect modification. All analyses were performed using R software (version 4.2.2). A two-sided *P* < 0.05 was considered statistically significant.

## Results

### Basic characteristics

Table 1 presents baseline characteristics of the 1,131 participants (65.7% women, mean age 69 years) by sex-stratified plasma ET quartiles. The median (Q1–Q3) ET concentrations for each quartile were 534.7 (430.6–613.0), 744.2 (693.3–819.6), 930.1 (861.1–1,007.2), and 1,200.0 (1,113.9–1,373.8) ng/mL, respectively.

**Table 1.** Baseline Characteristics of Participants Stratified by Sex-Specific Quartiles of Plasma ET Concentrations.

|  | Overall (n = 1131) | Q1 (n = 271) | Q2 (n = 280) | Q3 (n = 288) | Q4 (n = 292) | P-value |
| --- | --- | --- | --- | --- | --- | --- |
| Plasma ET, ng/mL* | 842.2 (657.8-1052.5) | 534.7 (430.6-613.0) | 744.2 (693.3-819.6) | 930.1 (861.1-1007.2) | 1200.0 (1113.9-1373.8) | <0.001 |
| Age, years, mean (SD) | 69.0 (7.5) | 69.7 (7.6) | 69.7 (7.8) | 68.3 (7.2) | 68.5 (7.2) | 0.028 |
| Women, n (%) | 743 (65.7) | 179 (66.1) | 187 (66.8) | 188 (65.3) | 189 (64.7) | 0.959 |
| Educational level, n (%) |  |  |  |  |  | 0.001 |
| Illiterate | 30 (2.7) | 15 (5.5) | 8 (2.9) | 6 (2.1) | 1 (0.3) |  |
| Primary school | 92 (8.1) | 29 (10.7) | 25 (8.9) | 17 (5.9) | 21 (7.2) |  |
| Middle school | 376 (33.2) | 90 (33.2) | 99 (35.4) | 106 (36.8) | 81 (27.7) |  |
| High school or above | 633 (56.0) | 137 (50.6) | 148 (52.9) | 159 (55.2) | 189 (64.7) |  |
| Ever smokers, n (%) | 211 (18.7) | 52 (19.2) | 53 (18.9) | 58 (20.1) | 48 (16.4) | 0.699 |
| Ever drinkers, n (%) | 160 (14.1) | 30 (11.1) | 43 (15.4) | 55 (19.1) | 32 (11.0) | 0.013 |
| Diet score, mean (SD) | 3.0 (0.6) | 3.0 (0.7) | 3.0 (0.6) | 3.0 (0.5) | 3.0 (0.6) | 0.671 |
| BMI, kg/m<sup>2</sup>, mean (SD) | 24.0 (3.3) | 23.8 (3.4) | 24.2 (3.5) | 24.3 (3.3) | 23.9 (3.0) | 0.186 |
| Hypertension, n (%) | 905 (80.0) | 225 (83.0) | 227 (81.1) | 225 (78.1) | 228 (78.1) | 0.383 |
| Hyperlipidemia, n (%) | 787 (69.6) | 172 (63.5) | 193 (68.9) | 206 (71.5) | 216 (74.0) | 0.046 |
| Diabetes, n (%) | 420 (37.1) | 98 (36.2) | 100 (35.7) | 119 (41.3) | 103 (35.3) | 0.400 |
| Cardiovascular disease, n (%) | 740 (65.4) | 185 (68.3) | 177 (63.2) | 186 (64.6) | 192 (65.8) | 0.642 |
| APOE genotype<sup>†</sup>, n (%) |  |  |  |  |  | 0.325 |
| ε4 carriers | 200 (17.7) | 56 (20.7) | 40 (14.3) | 54 (18.8) | 50 (17.1) |  |
| ε4 non-carriers | 907 (80.2) | 208 (76.8) | 237 (84.6) | 226 (78.5) | 236 (80.8) |  |
| ε2/ε4 carriers | 24 (2.1) | 7 (2.6) | 3 (1.1) | 8 (2.8) | 6 (2.1) |  |
| Baseline MoCA score, mean (SD) | 21.6 (4.3) | 21.1 (4.6) | 21.3 (4.6) | 21.8 (4.3) | 22.3 (3.7) | 0.002 |
| First follow-up MoCA score, mean (SD) | 21.4 (4.8) | 20.5 (4.9) | 20.7 (4.9) | 21.9 (4.9) | 22.5 (4.2) | <0.001 |
| Δ MoCA score, mean (SD) | -0.2 (3.4) | -0.6 (3.7) | -0.5 (3.3) | 0.1 (3.2) | 0.2 (3.3) | 0.008 |

Values are percentages for categorical variables and means ± SD for continuous variables. One-way ANOVA factor was used for continuous variables, and a $\chi^2$ test was used for categorical variables. $P < 0.05$ was considered significant.
\*Plasma ET concentrations are presented with the interquartile range (Q1–Q3).
<sup>†</sup>APOE ε4 carriers were defined as carriers of ε3/ε4 or ε4/ε4 genotypes.
BMI, Body Mass Index; APOE, Apolipoprotein E; MoCA, Montreal Cognitive Assessment.

Participants with higher ET levels were younger and demonstrated better educational attainment, and higher prevalence of hyperlipidemia.

Baseline MoCA scores were higher across ET quartiles (Q1:21.1±4.6, Q2:21.3±4.6, Q3:21.8±4.3, Q4:22.3±3.7; *P* = 0.002), and this pattern persisted at follow-up (Q1:20.5±4.9, Q2:20.7±4.9, Q3:21.9±4.9, Q4:22.5±4.2; *P* < 0.001). Change in total MoCA score (ΔMoCA) diverged significantly (*P* = 0.008), with declines observed in Q1 (−0.6±3.7) and Q2 (−0.5±3.3), contrasting with marginal improvements in Q3 (+0.1±3.2) and Q4 (+0.2±3.3).

### Assessment of potential confounding

Partial correlation analysis showed weak associations between ET and covariates, with all absolute values of partial correlation coefficients ≤0.15 (eFigure 2). In multivariable linear regression with ET as the outcome, female gender was positively associated with ET concentrations (*β*=0.39, 95% CI: 0.23–0.54; *P* < 0.001), as were higher educational attainment (high school or above vs. illiterate: *β*=0.46, 95% CI: 0.07–0.84; *P* = 0.02) and hyperlipidemia (*β*=0.20, 95% CI: 0.08–0.33; *P* = 0.001) (eFigure 3).

### Associations of baseline plasma ET levels with change in total MoCA scores

Associations between baseline plasma ET concentrations and change in total MoCA score are presented in Table 2. Each 1-SD increase in ET was positively associated with a 0.23 higher point in MoCA changes (*β*=0.23, 95% CI: 0.04–0.43; *P* = 0.021) after adjustment for age, sex, follow-up years, educational level, smoking, alcohol, diet score, BMI, hypertension, hyperlipidemia, diabetes, cardiovascular disease, *APOE ε4* status and baseline MoCA scores. The mean changes (95% CIs) in total MoCA score across ET quartiles were −0.58 (−0.74 to −0.43) for Q1, −0.52 (−0.68 to −0.37) for Q2, 0.09 (−0.07 to 0.24) for Q3, and 0.18 (0.03 to 0.33; *P* _for trend_ = 0.001) for Q4.

**Table 2.** Associations between Baseline Plasma ET Concentrations and Change in Total MoCA Score in the Total Population and Stratifying by Sex.

|  | ET (ng/mL) per 1-SD increment |  | Q1 | Q2 | Q3 | Q4 | *P* for trend |
| --- | --- | --- | --- | --- | --- | --- | --- |
| | $\beta$ (95% CI) | *P*-value | $\Delta$ MoCA score, mean (95% CI) | $\Delta$ MoCA score, mean (95% CI) | $\Delta$ MoCA score, mean (95% CI) | $\Delta$ MoCA score, mean (95% CI) | |
| Total |  |  |  |  |  |  |  |
| Model 1 | 0.19 (-0.02, 0.40) | 0.070 | -0.58 (-0.63, -0.54) | -0.52 (-0.57, -0.48) | 0.09 (0.04, 0.13) | 0.18 (0.14, 0.23) | 0.004 |
| Model 2 | 0.17 (-0.04, 0.38) | 0.114 | -0.58 (-0.65, -0.51) | -0.52 (-0.59, -0.46) | 0.09 (0.02, 0.15) | 0.18 (0.12, 0.24) | 0.006 |
| Model 3 | 0.23 (0.04, 0.43) | 0.021 | -0.58 (-0.74, -0.43) | -0.52 (-0.68, -0.37) | 0.09 (-0.07, 0.24) | 0.18 (0.03, 0.33) | 0.001 |
| Men |  |  |  |  |  |  |  |
| Model 1 | 0.75 (0.31, 1.19) | 0.001 | -1.33 (-1.63, -1.03) | -1.1 (-1.36, -0.84) | 0.36 (0.08, 0.64) | 0.35 (0.08, 0.61) | <0.001 |
| Model 2 | 0.79 (0.33, 1.24) | <0.001 | -1.33 (-1.52, -1.13) | -1.1 (-1.23, -0.96) | 0.36 (0.22, 0.50) | 0.35 (0.21, 0.49) | <0.001 |
| Model 3 | 0.78 (0.35, 1.21) | <0.001 | -1.33 (-1.38, -1.27) | -1.1 (-1.16, -1.04) | 0.36 (0.31, 0.41) | 0.35 (0.29, 0.40) | <0.001 |
| Women |  |  |  |  |  |  |  |
| Model 1 | 0.02 (-0.21, 0.25) | 0.842 | -0.20 (-0.40, 0) | -0.24 (-0.43, -0.05) | -0.06 (-0.25, 0.13) | 0.09 (-0.10, 0.28) | 0.468 |
| Model 2 | 0.02 (-0.26, 0.21) | 0.850 | -0.20 (-0.30, -0.10) | -0.24 (-0.34, -0.14) | -0.06 (-0.15, 0.03) | 0.09 (0.02, 0.16) | 0.732 |
| Model 3 | 0.06 (-0.16, 0.27) | 0.618 | -0.20 (-0.26, -0.14) | -0.24 (-0.30, -0.18) | -0.06 (-0.12, 0) | 0.09 (0.04, 0.14) | 0.346 |

Linear regression models were used: Model 1 was adjusted for age, sex and follow-up years. Model 2 was further adjusted for smoking, alcohol, diet score, educational level, BMI, *APOE* $\epsilon 4$ status, hypertension, hyperlipidemia, diabetes and cardiovascular disease. Model 3 was additionally adjusted for baseline MoCA scores.
The interaction between sex and continuous plasma ET was significant (LRT: $\chi^2$ (1) = 8.39, *P* = 0.004).
The interaction between sex and the four-category ET groups was also significant (LRT: $\chi^2$ (4) = 10.24, *P* = 0.017).
*P* for trend was tested in all models by treating ET categories as a continuous variable assigned the median value of each category.
Q1, quartile 1; Q2, quartile 2; Q3, quartile 3; Q4, quartile 4.

When analyses were stratified by sex, the positive association between plasma ET and MoCA score changes persisted only in men (*P* _for interaction_ = 0.004 for continuous ET; *P* _for interaction_ = 0.017 for ET quartiles). Among them, each 1-SD increase in ET was associated 0.78 higher total MoCA scores change (*β*=0.78, 95% CI: 0.35–1.21; *P* < 0.001). The mean changes (95% CIs) in total MoCA score across ET quartiles were −1.33 (−1.38 to −1.27) for Q1, −1.1 (−1.16 to −1.04) for Q2, 0.36 (0.31 to 0.41) for Q3 and 0.35 (0.29 to 0.40; *P* _for trend_ < 0.001) for Q4 among men. In contrast, no significant associations were observed in women.

In Figure 1, RCS showed no significant departure from linear associations overall (*P* _non-linearity_ = 0.143) or within sex strata (men: *P* _non-linearity_ = 0.577; women: *P* _non-linearity_ = 0.758). However, visual inspection of the spline curves revealed a potential plateauing effect at ET concentrations ≥1,000 ng/mL in the total population (Figure 1A). Baseline ET concentrations differed between men and women (eTable 1). Most men (81.5%) had concentrations below 1,000 ng/mL (median 754.2, IQR 592.0–937.9 ng/mL), while 35.7% of women exceeded 1,000 ng/mL (median 890.1, IQR 709.7–1,095.6 ng/mL, eFigure 4).

**Figure 1.**
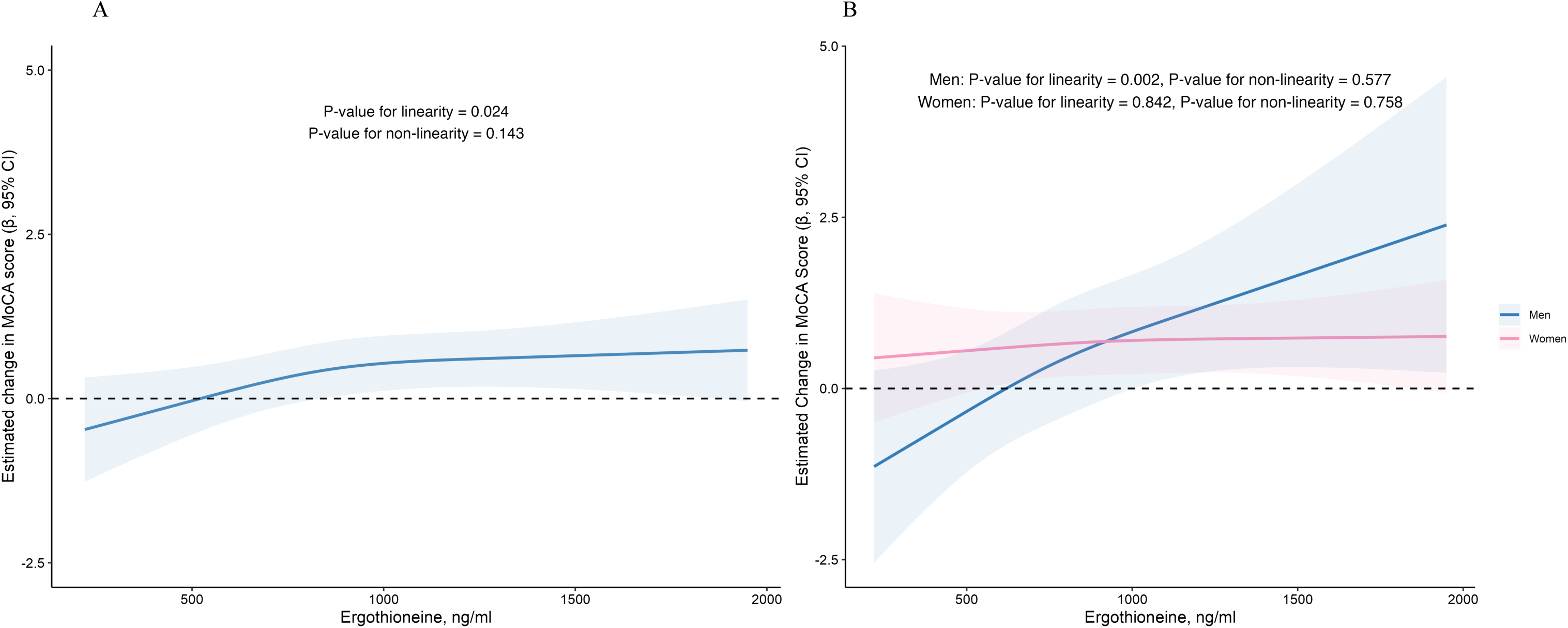
Multivariate-adjusted Relation of Baseline Plasma ET with MoCA Score Changes for the Total Population (A) and Stratifying by Sex (B). Associations were evaluated with the use of restricted cubic splines. The solid lines represent the central *β* estimate and the shaded area represent the 95% confidence intervals (CIs).

To facilitate clinical interpretation of effect sizes, we conducted an analysis substituting raw plasma ET concentrations (per 200 ng/mL increment) for z-scores in Model 3 (Figure 2). In the total cohort, each 200 ng/mL higher baseline ET was marginally associated with a 0.13-point improvement in MoCA change (*β*=0.13, 95% CI: 0.02–0.23), roughly equivalent to the association observed for 1.6 years younger age (*β*=0.08 per year decrease, 95% CI: 0.06–0.11). Among men, each 200 ng/mL higher ET was associated with a 0.43-point improvement in MoCA change (*β*=0.43, 95% CI: 0.19–0.66). This effect magnitude corresponds approximately to 4.3 years younger age, based on the age association (*β*=0.10 per year decrease, 95% CI: 0.05– 0.15). No significant associations were observed in women.

**Figure 2.**
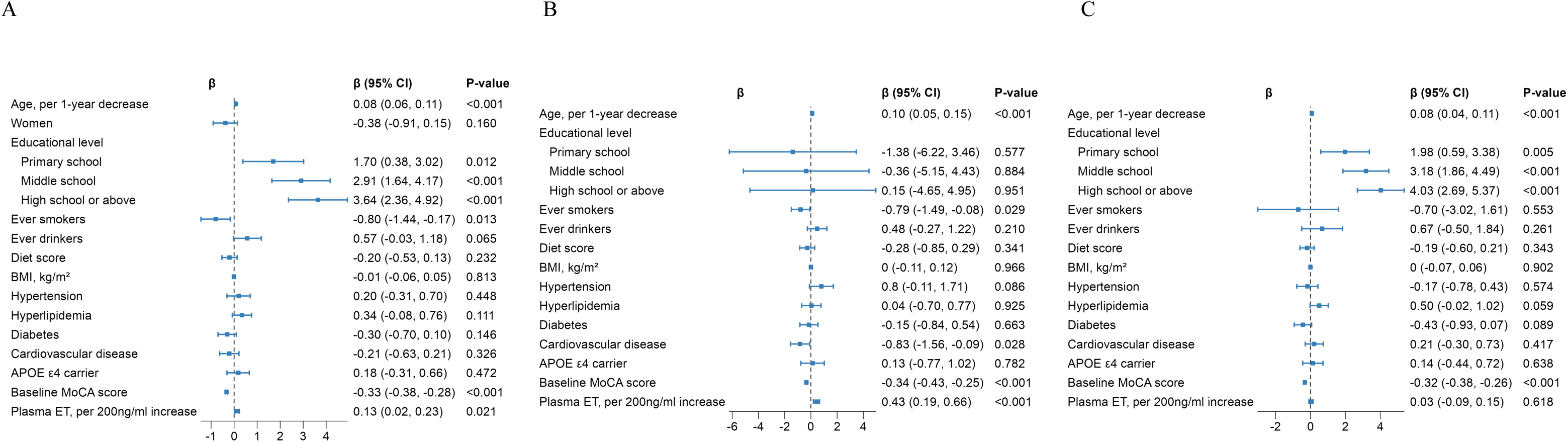
Forest Plots of Multivariable Associations Between Covariates and MoCA Score Change (A) Total population, (B) Men, (C) Women. Age analyzed per 1 year decrement. ET concentrations analyzed per 200 ng/mL increment.

### Plasma ET and specific cognitive domains of the MoCA score

Table 3 shows the association between baseline plasma ET concentrations and changes in each individual MoCA component. A significant positive association between ET (per 1 SD) and change in visuospatial/executive score (*β*=0.08, 95% CI: 0.02–0.14; *P* = 0.012) and delayed recall score (*β*=0.11, 95% CI: 0.02–0.20; *P* = 0.014) were observed after adjusting for potential confounders. Supplementary analyses characterizing the population and associations using the annualized change of total MoCA score (ΔMoCA / follow-up years) and MoCA domain score (ΔMoCA domain / follow-up years) are presented in eTables 2 and 3, which aligns with the main findings.

**Table 3.** Associations between Baseline Plasma ET Concentrations and Changes in MoCA Domain Scores in the Total Population.

|  | ET (ng/mL) per 1-SD increment |  | Q1 | Q2 | Q3 | Q4 | *P* for trend |
| --- | --- | --- | --- | --- | --- | --- | --- |
|  | *β* (95% CI) | *P*-value | Δ MoCA score, mean (95% CI) | Δ MoCA score, mean (95% CI) | Δ MoCA score, mean (95% CI) | Δ MoCA score, mean (95% CI) |  |
| **Visuospatial/executive** |  |  |  |  |  |  |  |
| Model 1 | 0.06 (-0.01, 0.13) | 0.085 | -0.17 (-0.18, -0.16) | -0.21 (-0.22, -0.20) | -0.13 (-0.14, -0.12) | 0.01 (0, 0.02) | 0.049 |
| Model 2 | 0.06 (-0.01, 0.13) | 0.115 | -0.17 (-0.19, -0.16) | -0.21 (-0.23, -0.19) | -0.13 (-0.15, -0.11) | 0.01 (-0.01, 0.03) | 0.061 |
| Model 3 | 0.08 (0.02, 0.14) | 0.012 | -0.17 (-0.24, -0.10) | -0.21 (-0.28, -0.15) | -0.13 (-0.20, -0.06) | 0.01 (-0.06, 0.08) | 0.016 |
| **Naming** |  |  |  |  |  |  |  |
| Model 1 | 0.01 (-0.03, 0.05) | 0.565 | 0.07 (0.07, 0.08) | -0.02 (-0.02, -0.02) | 0.03 (0.03, 0.03) | 0.08 (0.08, 0.08) | 0.622 |
| Model 2 | 0.01 (-0.04, 0.05) | 0.760 | 0.07 (0.06, 0.08) | -0.02 (-0.03, -0.01) | 0.03 (0.02, 0.04) | 0.08 (0.07, 0.09) | 0.828 |
| Model 3 | 0.02 (-0.02, 0.06) | 0.303 | 0.07 (0.03, 0.12) | -0.02 (-0.06, 0.01) | 0.03 (-0.01, 0.06) | 0.08 (0.04, 0.12) | 0.418 |
| **Attention** |  |  |  |  |  |  |  |
| Model 1 | -0.02 (-0.09, 0.05) | 0.621 | -0.15 (-0.15, -0.15) | -0.17 (-0.17, -0.17) | -0.02 (-0.03, -0.02) | -0.08 (-0.08, -0.07) | 0.277 |
| Model 2 | -0.03 (-0.10, 0.04) | 0.467 | -0.15 (-0.16, -0.13) | -0.17 (-0.18, -0.16) | -0.02 (-0.04, -0.01) | -0.08 (-0.09, -0.07) | 0.388 |
| Model 3 | 0 (-0.05, 0.06) | 0.890 | -0.15 (-0.22, -0.07) | -0.17 (-0.25, -0.09) | -0.02 (-0.09, 0.04) | -0.08 (-0.14, -0.01) | 0.081 |
| **Language** |  |  |  |  |  |  |  |
| Model 1 | 0.03 (-0.03, 0.08) | 0.310 | -0.03 (-0.04, -0.03) | -0.08 (-0.08, -0.07) | -0.01 (-0.01, 0) | 0.02 (0.02, 0.03) | 0.342 |
| Model 2 | 0.02 (-0.03, 0.08) | 0.371 | -0.03 (-0.05, -0.01) | -0.08 (-0.1, -0.06) | -0.01 (-0.02, 0.01) | 0.02 (0.01, 0.04) | 0.331 |
| Model 3 | 0.04 (-0.01, 0.08) | 0.120 | -0.03 (-0.09, 0.02) | -0.08 (-0.13, -0.02) | -0.01 (-0.06, 0.05) | 0.02 (-0.03, 0.08) | 0.179 |
| **Abstraction** |  |  |  |  |  |  |  |
| Model 1 | -0.02 (-0.08, 0.03) | 0.375 | -0.20 (-0.21, -0.19) | -0.03 (-0.04, -0.02) | -0.08 (-0.09, -0.07) | -0.09 (-0.10, -0.08) | 0.358 |
| Model 2 | -0.03 (-0.08, 0.03) | 0.356 | -0.20 (-0.21, -0.19) | -0.03 (-0.04, -0.02) | -0.08 (-0.09, -0.07) | -0.09 (-0.01, -0.08) | 0.364 |
| Model 3 | 0.01 (-0.04, 0.05) | 0.775 | -0.20 (-0.26, -0.14) | -0.03 (-0.09, 0.03) | -0.08 (-0.14, -0.02) | -0.09 (-0.15, -0.03) | 0.085 |
| **Delayed recall** |  |  |  |  |  |  |  |
| Model 1 | 0.10 (-0.01, 0.21) | 0.064 | 0.06 (0.05, 0.08) | 0.24 (0.22, 0.25) | 0.43 (0.42, 0.44) | 0.38 (0.36, 0.39) | 0.032 |
| Model 2 | 0.11 (0, 0.21) | 0.056 | 0.06 (0.03, 0.09) | 0.24 (0.21, 0.26) | 0.43 (0.40, 0.46) | 0.38 (0.35, 0.40) | 0.029 |
| Model 3 | 0.11 (0.02, 0.20) | 0.014 | 0.06 (-0.05, 0.18) | 0.24 (0.12, 0.35) | 0.43 (0.31, 0.55) | 0.38 (0.26, 0.49) | 0.004 |
| **Orientation** |  |  |  |  |  |  |  |
| Model 1 | 0.03 (-0.02, 0.08) | 0.250 | -0.17 (-0.17, -0.16) | -0.25 (-0.25, -0.24) | -0.13 (-0.14, -0.13) | -0.14 (-0.15, -0.13) | 0.479 |
| Model 2 | 0.02 (-0.02, 0.07) | 0.331 | -0.17 (-0.18, -0.15) | -0.25 (-0.26, -0.24) | -0.13 (-0.14, -0.12) | -0.14 (-0.15, -0.13) | 0.601 |
| Model 3 | 0.02 (-0.02, 0.06) | 0.396 | -0.17 (-0.22, -0.11) | -0.25 (-0.29, -0.2) | -0.13 (-0.18, -0.09) | -0.14 (-0.18, -0.10) | 0.412 |

*P* for trend was tested in all models by treating ET categories as a continuous variable assigned the median value of each category. Q, quartile.

### Stratified analysis

No statistically significant interactions were detected across any subgroups (eFigure 5A-D; all *P* _for interaction_ > 0.05), yet the direction and magnitude of associations between baseline plasma ET and MoCA score change exhibited heterogeneity in specific strata.

## Discussion

In this prospective cohort study of 1,131 community-dwelling older adults (mean age 69 years), higher baseline plasma ET levels were significantly associated with slower cognitive decline, as assessed by MoCA scores, during a 2-year follow-up period.

More specifically, when the plasma concentration of ET exceeds 1,000 ng/mL, the decline in cognitive function significantly slows down. However, this association has only been observed in men. Domain-specific analysis found that the observed ET-MoCA association was mainly driven by the temporary slowdown in the decline of visuospatial/executive and delayed recall.

While cross-sectional studies have consistently shown that higher ET levels were significantly associated with higher cognitive function^12,13^ and lower risk of dementia^14–17^, longitudinal evidence remains scarce and methodologically limited. A previous French study on the basis of a nested case-control study design (N=842) screened the associations of over 10 food metabolites with cognitive function and found that higher ET level was significantly associated with slower cognitive decline^25^. Similarly, a study from Singapore^18^ reported that lower ET level was associated with accelerated cognitive decline among 470 elderly participants, yet the sample of this study could not represent the general community-dwelling elderly, as 40% of the participants had been diagnosed with dementia at baseline. From the perspective of dementia prevention, exploring the association between plasma ET and cognitive function in a community-dwelling elderly population is of greater public health significance, and our study has filled this gap. Notably, determining the temporal sequence between the decline in ET levels and cognitive decline is crucial, as it is possible that cognitive impairment directly affects dietary intake, leading to reduced consumption of ET-rich foods and a decline in plasma ET levels. Previous studies have struggled to address this question as they did not exclude participants with dementia at baseline^18^. In contrast, our study included only participants with normal cognitive function, and the results remained robust even after excluding those with baseline cognitive function at the lower end of the normal range. Collectively, our findings support that low ET intake occurs prior to cognitive decline.

The observed associations between ET levels and specific cognitive domains, namely delayed recall and visuospatial/executive function, warrant consideration regarding ET’s potential neuroprotective roles. Impaired delayed recall represents one of the earliest and most sensitive cognitive markers of dementia progression, predictive of conversion from MCI to dementia^26,27^. The preferential preservation of this function by ET suggests targeted neuroprotective effects within the hippocampus, the brain region orchestrating declarative memory consolidation and retrieval.

Experimental evidence shows that ET protects hippocampal neurons from oxidative stress^28^, reduces Aβ accumulation and lipid peroxidation^29^ in the hippocampus, and ET deficiency disrupts hippocampal neurogenesis^30^ while supplementation promotes dendritic spine formation^31^. Human neuroimaging studies corroborate this hippocampal vulnerability, showing reduced hippocampal volume in MCI and dementia patients with systemic ET deficiency^14,18^. Specifically, visuospatial/executive function critically depends on the prefrontal cortex^32^ and fronto-parietal network modulation. The susceptibility of these regions to oxidative stress and metabolic dysregulation suggests they might be sensitive to ET’s antioxidant properties, though the precise mechanisms require further exploration.

The men-specific cognitive changes associated with ET likely reflect a dose-dependent association modulated by baseline sex differences. Women exhibited substantially higher median plasma ET concentrations (890.1 ng/mL; IQR: 709.7– 1,095.6) than men (754.2 ng/mL; IQR: 592.0–937.9), with 35.7% of women exceeding the 1,000 ng/mL saturation threshold where dose-response relationships plateaued in our analysis. Conversely, most men (81.5%) operated below this threshold, possibly enabling detectable dose-dependent neuroprotection. This aligns with broader nutrient-sex interactions wherein antioxidants (e.g., vitamin C^33^, fish-oil^34^) show men-predominant associations for cognitive function, possibly due to women’s higher baseline antioxidant reserves. Human microbiome studies demonstrated divergent bacterial communities between sexes^35^, the sexual dimorphism in microbiome could theoretically modulate ET bioavailability through bacterial absorption^36–38^ or degradation^39,40^, although direct evidence is lacking.

Ultimately, the dose effect observed for ET provides the most plausible explanation for this sex divergence, aligning with plateau effects observed for other neuroprotective agents and consistent with our cohort’s ET distribution. However, the potential contribution of alternative mechanisms requires further investigation.

Our study design and analytical approaches robustly mitigate concerns regarding reverse causation. First, the longitudinal analysis of baseline plasma ET levels against subsequent changes in cognitive function inherently establishes temporal precedence. Second, the positive association between ET and MoCA score changes strengthened rather than attenuated after comprehensive adjustment for baseline MoCA scores in multivariable Model 3, contradicting the expectation that confounding by pre-existing cognitive status would diminish the effect. Third, weak correlations (|r| ≤0.15) between baseline ET levels and key covariates—including demographics, lifestyle factors, and comorbidities—indicate minimal confounding potential, a conclusion further reinforced by our inclusion of these variables in all primary models.

There are also limitations to our study. First, most variables, including demographic data, health conditions and diet were self-reported, which could be inherently challenging in elderly populations due to recall bias and cognitive limitations. Second, the median 2-year follow-up period, though sufficient to detect significant cognitive changes in this cohort, precluded the observation of long-term associations. Extended surveillance would clarify ET’s durability in dementia prevention. Third, reliance ET was only measured at baseline therefore cannot capture potential intraindividual fluctuations over time, although the observed plateau effect above 1,000 ng/mL suggests stable biological activity beyond this threshold. Fourth, the difference of standardized experimental and analytical procedures in metabolomics (e.g., instrumentation, quantification methods, sample preparation) means that the ET concentrations quantified in this study cannot be directly compared with quantitative results from other metabolomic investigations. Finally, as participants were exclusively community-dwelling Chinese older adults, generalizability to younger populations, other ethnicities, or clinical cohorts requires verification. Notwithstanding these limitations, our use of a quantitative biomarker, prospective design, comprehensive confounder adjustment, and stratification analyses provides robust evidence supporting a sex-specific protective role of ET against cognitive decline in older adults.

In conclusion, our findings indicate that higher plasma ET levels are significantly associated with slower cognitive decline independent of confounders in general community elderly participants without dementia at baseline, whereas such association was only observed in men but not in women. Further analysis with large sample size and longer follow-up period is needed to verify our findings.

## Article information

### Author Contributions

Xiao and Zong had full access to all of the data in the study and takes responsibility for the integrity of the data and the accuracy of the data analysis.

*Concept and design:* Zhao, Xiao and Zong.

*Acquisition, analysis, or interpretation of data:* All authors.

*Drafting of the manuscript:* Zhao, Zong.

*Critical revision of the manuscript for important intellectual content:* All authors.

*Statistical analysis:* Zhao, Feng, Li and Liu.

*Administrative, technical, or material support:* Lu, Jiang and Xiao.

*Supervision:* Xiao and Zong.

### Additional Contributions

This study was conducted using the Shanghai Brain Aging Study. All authors are thankful to the participants agreed to contribute to study. We also gratefully acknowledge Xiamen Kingdomway Group Company for their in-kind contribution of the ergothioneine reference standard used in this study.

## Data Availability

All data produced in the present study are available upon reasonable request to the authors

