## supplemental files for "Associations of plasma ergothioneine levels with cognitive function change in non-demented older Chinese adults: A community-based longitudinal study"

**Supplementary Figures**

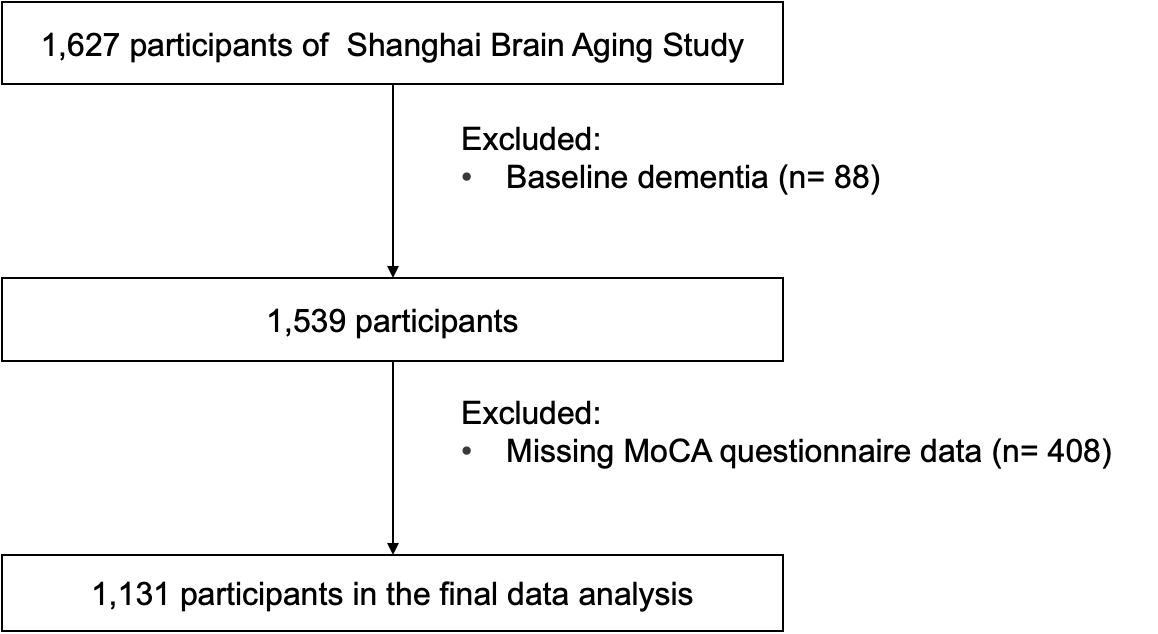

eFigure 1. Flow diagram of participants in Shanghai Brain Aging Study. Dementia cases at baseline were ascertained through a comprehensive approach incorporating standardized cognitive screening, confirmatory neuropsychological evaluation, and verification via health insurance data.

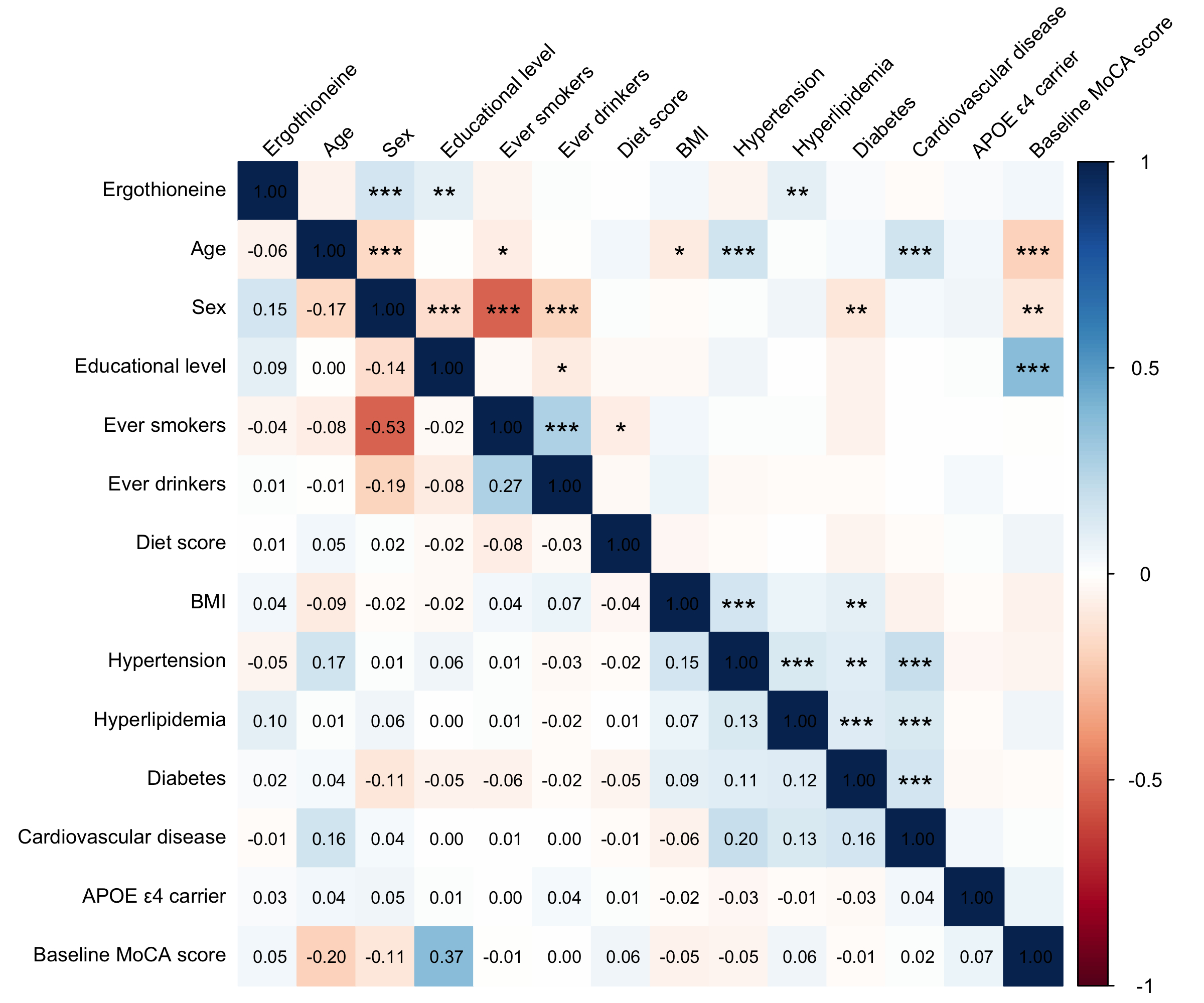

eFigure 2. Heatmap of Partial Correlations between Plasma ET and Covariates. Blue/red shades indicate positive/negative correlations, with asterisks denoting statistical significance (*P <0.05, **P <0.01，***P <0.001)

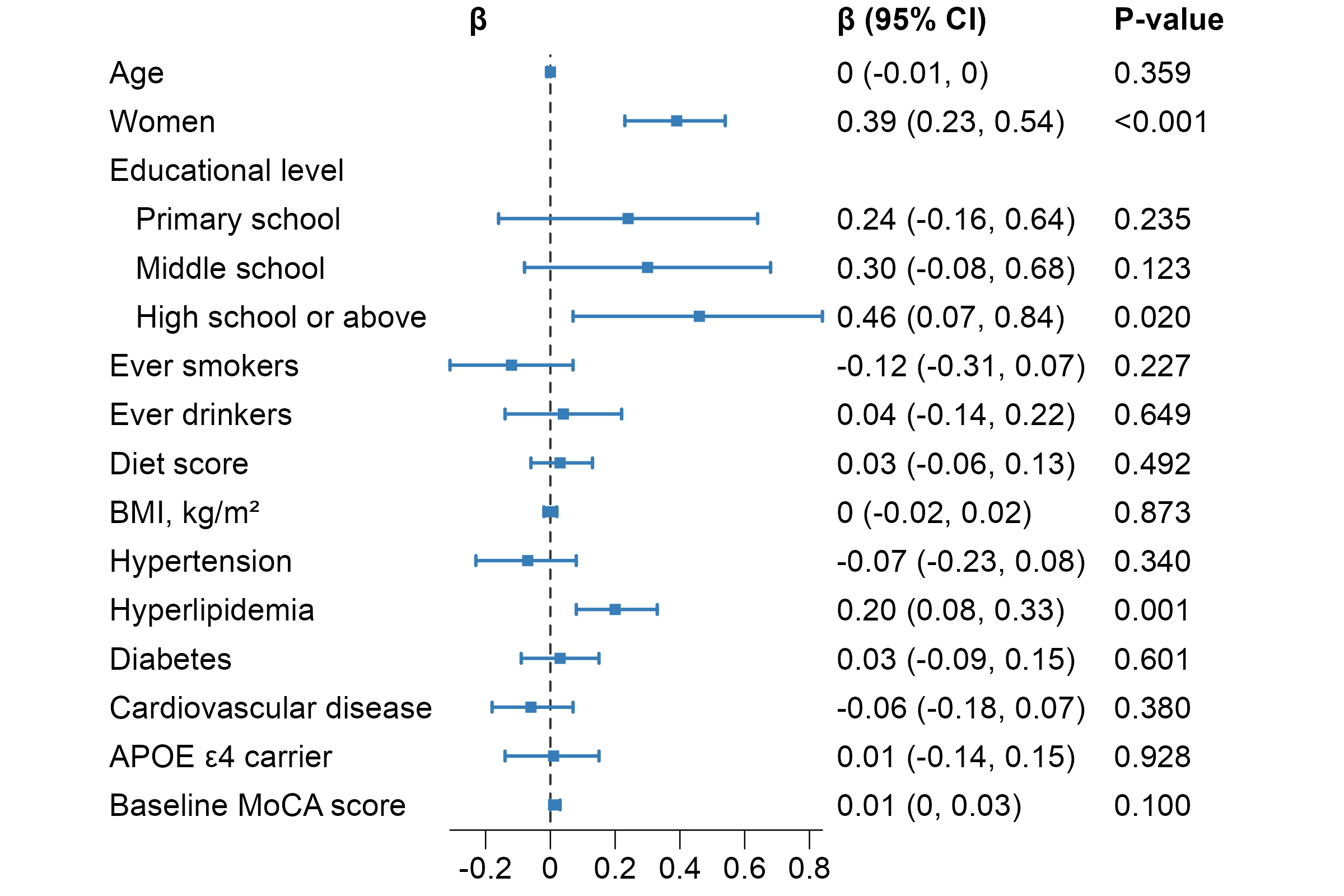

eFigure 3. Forest Plot of Multivariable Linear Regression Coefficients (β) for Factors Associated with Baseline Plasma ET Levels. Error bars represent 95% confidence intervals.

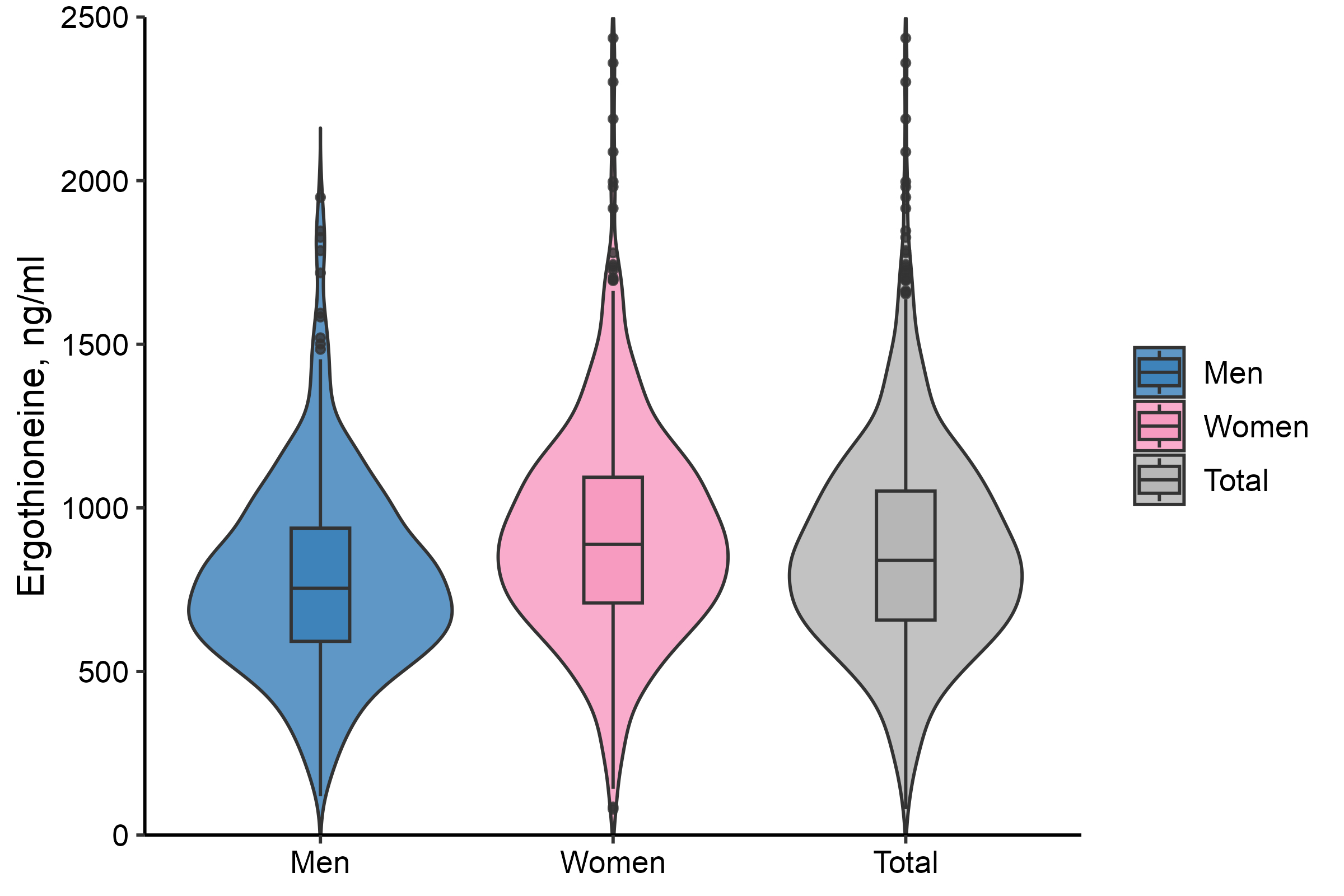

eFigure 4. Distribution of Baseline Plasma ET Concentrations (ng/mL) in the Total Population and by Sex. Violin-box plots display the probability density distribution (violin shape) with overlaid box plots showing the median (central line), interquartile range (IQR; box boundaries), and whiskers extending to 1.5×IQR. The y-axis is truncated to 0-2500 ng/mL for enhanced visualization of the central distribution, while the full data range spans 79.1-4356.5 ng/mL (outliers retained in analysis).

A	B

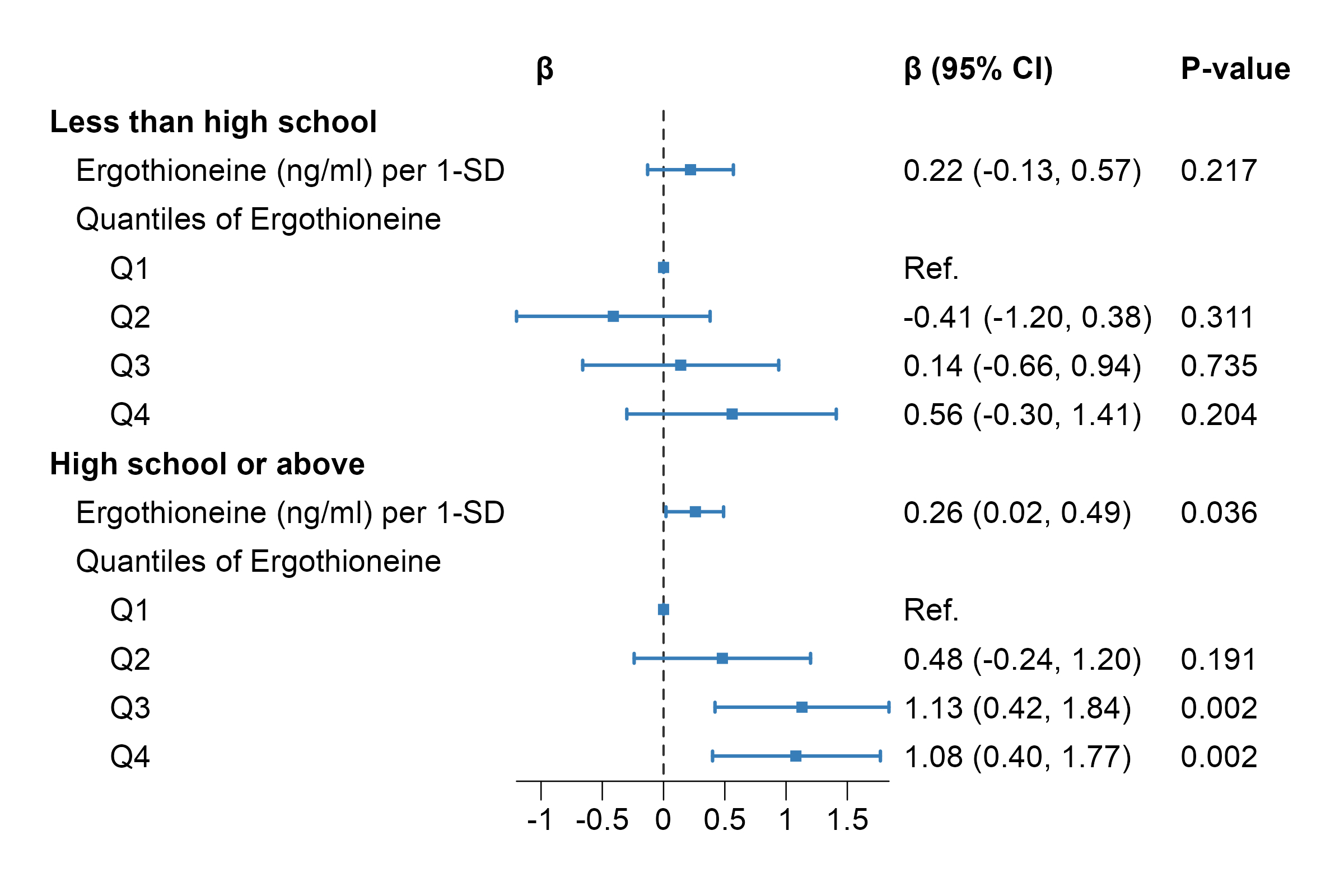

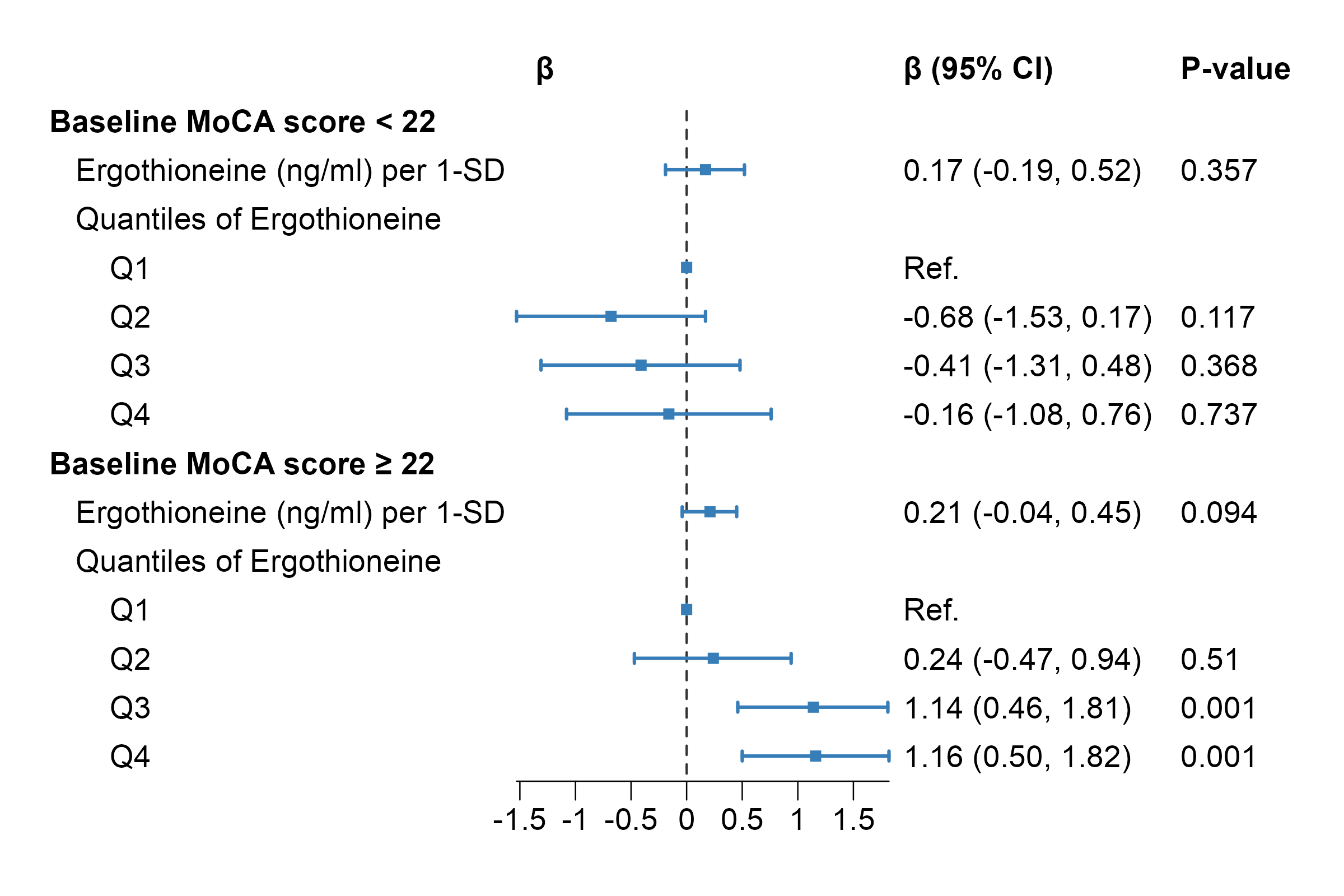

C	D

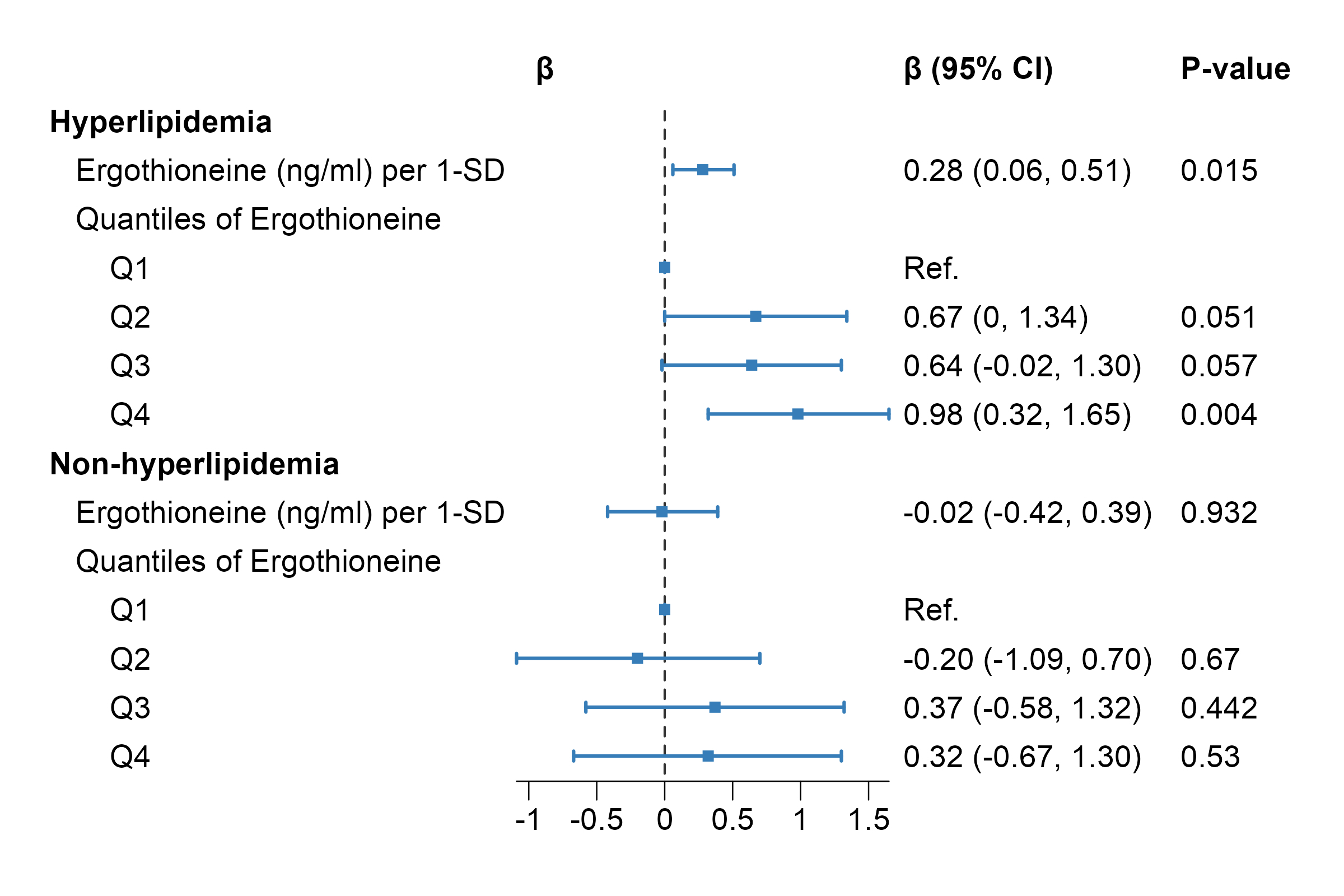

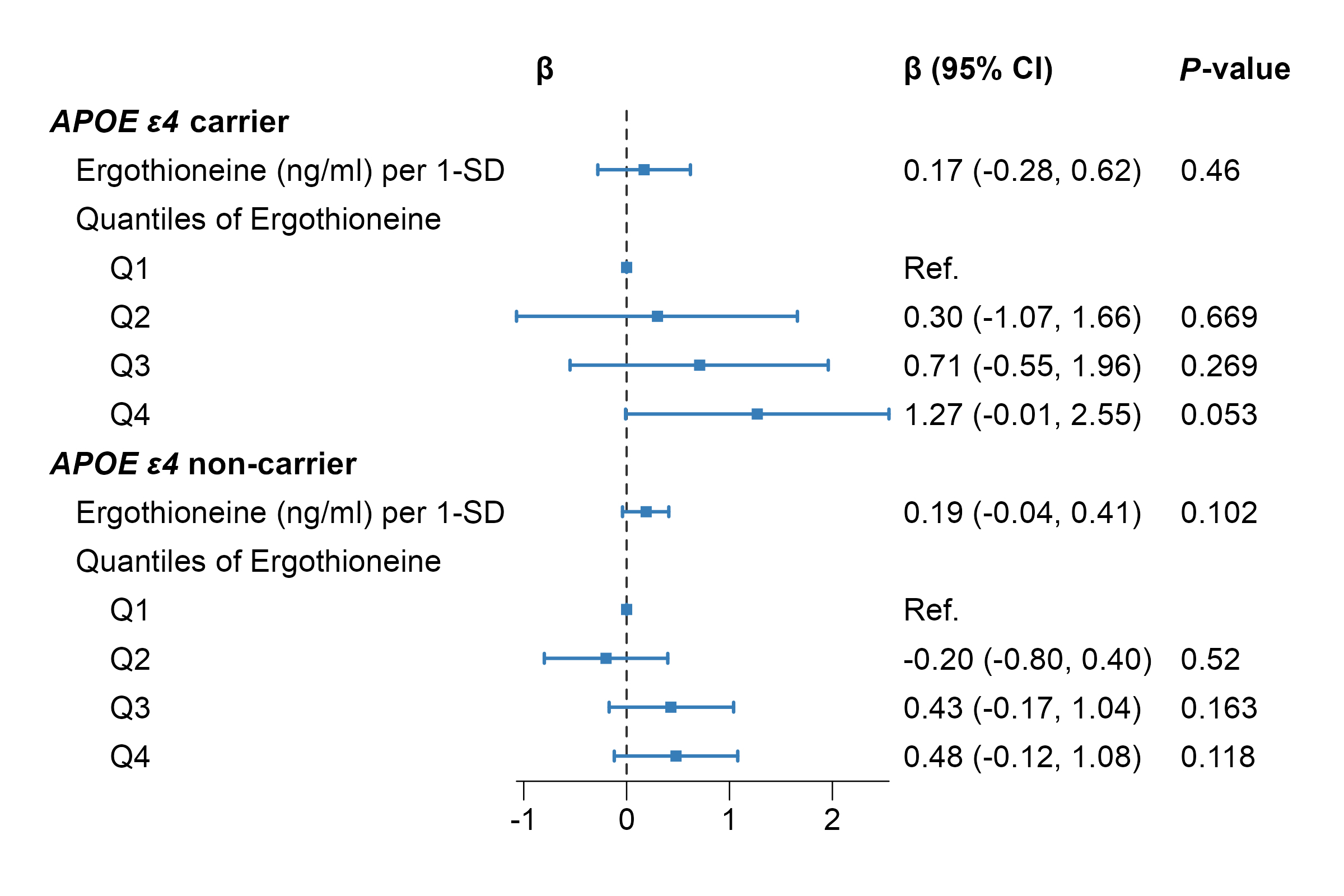

eFigure 5. Adjusted Association of Plasma ET with MoCA Score Changes by Educational level (A), Baseline MoCA Score Groups (B), Hyperlipidemia Status (C) and

*APOE* genotype (D). The interaction between educational levels and continue ET was not significant (LRT: χ² (1) = 0.01, *P* = 0.93). The interaction between baseline MoCA groups and continue ET was not significant (LRT: χ² (1) = 0.10, *P* = 0.75). The interaction between hyperlipidemia status and continue ET was not significant (LRT: χ² (1) = 1.53, *P* = 0.22). The interaction between *APOE* genotype and continue ET was not significant (LRT: χ² (1) = 1.03, *P* = 0.59).

**Supplementary Tables**

**eTable 1. General Characteristics of the Study Population at Baseline by Sex**

|  | Overall (n = 1131) | Men (n = 388) | Women (n = 743) | *P*-value |
| --- | --- | --- | --- | --- |
| Plasma ET^*^, ng/mL | 842.2 (657.8-1052.5) | 754.2 (592.0–937.9) | 890.1 (709.7–1095.6) | <0.001 |
| Age, years, mean (SD) | 69.0 (7.5) | 70.3 (7.3) | 68.4 (7.5) | <0.001 |
| Educational level, n (%) |  |  |  | <0.001 |
| Illiterate | 30 (2.7) | 2 (0.5) | 28 (3.8) |  |
| Primary school | 92 (8.1) | 28 (7.2) | 64 (8.6) |  |
| Middle school | 376 (33.2) | 101 (26.0) | 275 (37.0) |  |
| High school or above | 633 (56.0) | 257 (66.2) | 376 (50.6) |  |
| Ever smokers, n (%) | 211 (18.7) | 204 (52.6) | 7 (0.9) | <0.001 |
| Ever drinkers, n (%) | 160 (14.1) | 132 (34.0) | 28 (3.8) | <0.001 |
| Diet score, mean (SD) | 3.0 (0.6) | 2.9 (0.6) | 3.0 (0.6) | 0.001 |
| BMI, kg/m^2^, mean (SD) | 24.0 (3.3) | 24.2 (3.0) | 23.9 (3.4) | 0.155 |
| Hypertension, n (%) | 905 (80.0) | 316 (81.4) | 589 (79.3) | 0.431 |
| Hyperlipidemia, n (%) | 787 (69.6) | 253 (65.2) | 534 (71.9) | 0.025 |
| Diabetes, n (%) | 420 (37.1) | 165 (42.5) | 255 (34.3) | 0.008 |
| Cardiovascular disease, n (%) | 740 (65.4) | 251 (64.7) | 489 (65.8) | 0.756 |
| *APOE* genotype^†^, n (%) |  |  |  | 0.294 |
| *ε4* carriers | 200 (17.7) | 65 (16.8) | 135 (18.2) |  |
| *ε4* non-carriers | 907 (80.2) | 318 (82.0) | 589 (79.3) |  |
| *ε2/ε4* carriers | 24 (2.1) | 5 (1.3) | 19 (2.6) |  |
| Baseline MoCA score, mean (SD) | 21.6 (4.3) | 22.4 (4.0) | 21.2 (4.4) | <0.001 |
| Follow-up MoCA score, mean (SD) | 21.4 (4.8) | 22.0 (4.6) | 21.1 (4.9) | 0.004 |
| Δ MoCA score, mean (SD) | -0.2 (3.4) | -0.4 (3.5) | -0.1 (3.3) | 0.170 |

Values are percentages for categorical variables and means ± SD for continuous variables. One-way ANOVA factor was used for continuous variables, and a χ2 test was used for categorical variables. *P* < 0.05 was considered significant.

*Plasma ET concentrations are presented with the interquartile range (Q1–Q3).

†*APOE ε4* carriers were defined as carriers of *ε3/ε4* or *ε4/ε4* genotypes.

BMI, Body Mass Index; APOE, Apolipoprotein E; MoCA, Montreal Cognitive Assessment.

eTable 2. Associations between Baseline Plasma ET Concentrations and Annualized Change in Total MoCA Score in the Total Population and Stratifying by Sex

|  | ET (ng/mL) per 1-SD increment | |  | Q1 |  | Q2 |  | Q3 |  | Q4 | *P* for trend |
| --- | --- | --- | --- | --- | --- | --- | --- | --- | --- | --- | --- |
|  | *β* (95% CI) | *P-*value |  | Annualized Δ MoCA, mean (95% CI) |  | Annualized Δ MoCA, mean (95% CI) |  | Annualized Δ MoCA, mean (95% CI) |  | Annualized Δ MoCA, mean (95% CI) |  |
| Total |  |  |  |  |  |  |  |  |  |  |  |
| Model 1 | 0.10 (0, 0.20) | 0.056 |  | -0.30 (-0.33, -0.28) |  | -0.27 (-0.29, -0.24) |  | 0.05 (0.03, 0.07) |  | 0.09 (0.07, 0.11) | 0.003 |
| Model 2 | 0.09 (-0.02, 0.19) | 0.095 |  | -0.30 (-0.34, -0.27) |  | -0.27 (-0.30, -0.23) |  | 0.05 (0.02, 0.08) |  | 0.09 (0.06, 0.12) | 0.005 |
| Model 3 | 0.12 (0.02, 0.22) | 0.016 |  | -0.30 (-0.38, -0.23) |  | -0.27 (-0.34, -0.19) |  | 0.05 (-0.03, 0.13) |  | 0.09 (0.02, 0.17) | 0.001 |
| Men |  |  |  |  |  |  |  |  |  |  |  |
| Model 1 | 0.38 (0.16, 0.61) | 0.001 |  | -0.68 (-0.71, -0.65) |  | -0.55 (-0.58, -0.52) |  | 0.19 (0.17, 0.22) |  | 0.17 (0.15, 0.20) | <0.001 |
| Model 2 | 0.40 (0.17, 0.63) | 0.001 |  | -0.68 (-0.83, -0.53) |  | -0.55 (-0.68, -0.42) |  | 0.19 (0.05, 0.34) |  | 0.17 (0.04, 0.30) | <0.001 |
| Model 3 | 0.40 (0.18, 0.61) | <0.001 |  | -0.68 (-0.83, -0.53) |  | -0.55 (-0.68, -0.42) |  | 0.19 (0.05, 0.34) |  | 0.17 (0.04, 0.30) | <0.001 |
| Women |  |  |  |  |  |  |  |  |  |  |  |
| Model 1 | 0.02 (-0.10, 0.13) | 0.791 |  | -0.11 (-0.14, -0.08) |  | -0.13 (-0.16, -0.10) |  | -0.03 (-0.06, 0) |  | 0.05 (0.02, 0.07) | 0.420 |
| Model 2 | -0.01 (-0.12, 0.11) | 0.896 |  | -0.11 (-0.16, -0.06) |  | -0.13 (-0.18, -0.08) |  | -0.03 (-0.07, 0.01) |  | 0.05 (0.01, 0.08) | 0.678 |
| Model 3 | 0.03 (-0.08, 0.14) | 0.575 |  | -0.11 (-0.21, -0.01) |  | -0.13 (-0.22, -0.03) |  | -0.03 (-0.13, 0.07) |  | 0.05 (-0.05, 0.14) | 0.307 |

Annualized change in total MoCA score = (MoCA _follow-up1_ –MoCA _baseline_) / follow-up years.

Linear regression models were used: Model 1 was adjusted for age and sex. Model 2 was further adjusted for smoking, alcohol, diet score, educational level, BMI, *APOE ε4* status, hypertension, hyperlipidemia, diabetes and cardiovascular disease. Model 3 was additionally adjusted for baseline MoCA scores.

The interaction between sex and continuous plasma ET was significant (LRT: χ² (1) = 8.64, *P* = 0.003).

The interaction between sex and the four-category ET groups was also significant (LRT: χ² (4) = 10.65, *P* = 0.014).

*P* for trend was tested in all models by treating ET categories as a continuous variable assigned the median value of each category.

Q1, quartile 1; Q2, quartile 2; Q3, quartile 3; Q4, quartile 4.

eTable 3. Associations between Baseline Plasma ET Concentrations and Annualized Change in MoCA domain Scores in the Total Population

|  | ET (ng/mL) per 1-SD increment | |  | | Q1 |  | Q2 |  | Q3 |  | Q4 | *P* for trend |
| --- | --- | --- | --- | --- | --- | --- | --- | --- | --- | --- | --- | --- |
|  | *β* (95% CI) | *P-*value |  | Annualized Δ MoCA, mean (95% CI) | |  | Δ MoCA score, mean (95% CI) |  | Δ MoCA score, mean (95% CI) |  | Δ MoCA score, mean (95% CI) |  |
| **Visuospatial/executive** |  |  |  |  | |  |  |  |  |  |  |  |
| Model 1 | 0.03 (0, 0.07) | 0.062 |  | -0.09 (-0.10, -0.09) | |  | -0.11 (-0.11, -0.10) |  | -0.07 (-0.07, -0.06) |  | 0.01 (0, 0.01) | 0.031 |
| Model 2 | 0.03 (0, 0.07) | 0.091 |  | -0.09 (-0.10, -0.09) | |  | -0.11 (-0.12, -0.10) |  | -0.07 (-0.07, -0.06) |  | 0.01 (0, 0.02) | 0.042 |
| Model 3 | 0.04 (0.01, 0.07) | 0.008 |  | -0.09 (-0.13, -0.06) | |  | -0.11 (-0.14, -0.07) |  | -0.07 (-0.10, -0.03) |  | 0.01 (-0.03, 0.04) | 0.010 |
| **Naming** |  |  |  |  | |  |  |  |  |  |  |  |
| Model 1 | 0.01 (-0.01, 0.03) | 0.581 |  | 0.04 (0.04, 0.04) | |  | -0.01 (-0.01, -0.01) |  | 0.01 (0.01, 0.02) |  | 0.04 (0.04, 0.04) | 0.645 |
| Model 2 | 0 (-0.02, 0.02) | 0.760 |  | 0.04 (0.03, 0.04) | |  | -0.01 (-0.01, 0) |  | 0.01 (0.01, 0.02) |  | 0.04 (0.04, 0.04) | 0.838 |
| Model 3 | 0.01 (-0.01, 0.03) | 0.308 |  | 0.04 (0.02, 0.06) | |  | -0.01 (-0.03, 0.01) |  | 0.01 (0, 0.03) |  | 0.04 (0.02, 0.06) | 0.432 |
| **Attention** |  |  |  |  | |  |  |  |  |  |  |  |
| Model 1 | -0.01 (-0.04, 0.03) | 0.673 |  | -0.07 (-0.07, -0.07) | |  | -0.09 (-0.09, -0.09) |  | -0.01 (-0.01, -0.01) |  | -0.04 (-0.04, -0.03) | 0.256 |
| Model 2 | -0.01 (-0.05, 0.02) | 0.513 |  | -0.07 (-0.08, -0.07) | |  | -0.09 (-0.09, -0.08) |  | -0.01 (-0.02, -0.01) |  | -0.04 (-0.04, -0.03) | 0.358 |
| Model 3 | 0 (-0.03, 0.03) | 0.825 |  | -0.07 (-0.11, -0.03) | |  | -0.09 (-0.13, -0.05) |  | -0.01 (-0.05, 0.02) |  | -0.04 (-0.07, 0) | 0.071 |
| **Language** |  |  |  |  | |  |  |  |  |  |  |  |
| Model 1 | 0.01 (-0.01, 0.04) | 0.284 |  | -0.02 (-0.02, -0.01) | |  | -0.04 (-0.04, -0.04) |  | 0 (-0.01, 0) |  | 0.01 (0.01, 0.02) | 0.303 |
| Model 2 | 0.01 (-0.01, 0.04) | 0.351 |  | -0.02 (-0.03, -0.01) | |  | -0.04 (-0.05, -0.03) |  | 0 (-0.01, 0.01) |  | 0.01 (0.01, 0.02) | 0.302 |
| Model 3 | 0.02 (0, 0.04) | 0.109 |  | -0.02 (-0.05, 0.01) | |  | -0.04 (-0.07, -0.01) |  | 0 (-0.03, 0.03) |  | 0.01 (-0.01, 0.04) | 0.157 |
| **Abstraction** |  |  |  |  | |  |  |  |  |  |  |  |
| Model 1 | -0.01 (-0.04, 0.01) | 0.384 |  | -0.10 (-0.11, -0.10) | |  | -0.01 (-0.02, -0.01) |  | -0.04 (-0.04, -0.04) |  | -0.05 (-0.05, -0.04) | 0.342 |
| Model 2 | -0.01 (-0.04, 0.01) | 0.377 |  | -0.10 (-0.11, -0.10) | |  | -0.01 (-0.02, -0.01) |  | -0.04 (-0.05, -0.03) |  | -0.05 (-0.05, -0.04) | 0.340 |
| Model 3 | 0 (-0.02, 0.03) | 0.739 |  | -0.10 (-0.13, -0.07) | |  | -0.01 (-0.05, 0.02) |  | -0.04 (-0.07, -0.01) |  | -0.05 (-0.08, -0.02) | 0.076 |
| **Delayed recall** |  |  |  |  | |  |  |  |  |  |  |  |
| Model 1 | 0.05 (0, 0.10) | 0.062 |  | 0.03 (0.02, 0.03) | |  | 0.11 (0.10, 0.12) |  | 0.22 (0.21, 0.23) |  | 0.18 (0.17, 0.19) | 0.031 |
| Model 2 | 0.05 (0, 0.11) | 0.055 |  | 0.03 (0.01, 0.04) | |  | 0.11 (0.10, 0.12) |  | 0.22 (0.21, 0.23) |  | 0.18 (0.17, 0.19) | 0.029 |
| Model 3 | 0.06 (0.01, 0.10) | 0.014 |  | 0.03 (-0.03, 0.08) | |  | 0.11 (0.05, 0.17) |  | 0.22 (0.16, 0.28) |  | 0.18 (0.12, 0.24) | 0.005 |
| **Orientation** |  |  |  |  | |  |  |  |  |  |  |  |
| Model 1 | 0.01 (-0.01, 0.04) | 0.253 |  | -0.08 (-0.09, -0.08) | |  | -0.12 (-0.12, -0.11) |  | -0.07 (-0.07, -0.06) |  | -0.07 (-0.07, -0.07) | 0.482 |
| Model 2 | 0.01 (-0.01, 0.04) | 0.325 |  | -0.08 (-0.09, -0.08) | |  | -0.12 (-0.12, -0.11) |  | -0.07 (-0.07, -0.06) |  | -0.07 (-0.07, -0.06) | 0.590 |
| Model 3 | 0.01 (-0.01, 0.03) | 0.389 |  | -0.08 (-0.11, -0.06) | |  | -0.12 (-0.14, -0.1) |  | -0.07 (-0.09, -0.04) |  | -0.07 (-0.09, -0.05) | 0.398 |

Annualized change in MoCA domain score = (MoCA domain _follow-up1_ –MoCA domain _baseline_) / follow-up years. Linear regression models were used. Adjustment for covariates in Models 1 and 2 was identical to eTable 2. Model 3 was additionally adjusted for baseline MoCA domain scores. *P* for trend was tested using the same method as in eTable 2. Q, quartile.
